# Comparing Missing Data Imputation Methods for Patient-Reported Outcomes in Esophageal Cancer Research

**DOI:** 10.1101/2025.09.10.25335531

**Authors:** Yong Jin Kweon, Emad A. Mohammed, Yousif Salman, Shayan Dhillon, Sara Najmeh, Carmen Mueller, Jonathan Cools-Lartigue, Jonathan Spicer, Lorenzo Ferri, Mehrnoush Dehghani, R. Trafford Crump

## Abstract

Missing data is common in patient-reported outcomes (PRO) research, particularly in oncology settings. We evaluated common methods for handling missing data in esophageal cancer quality of life measurements, namely: multiple imputation by chained equations, variational autoencoder, denoising autoencoder, Bayesian principal component analysis, a deep autoen-coder method with patient-specific embeddings and temporal pattern modeling, SoftImpute, and K-nearest neighbors. Using data from McGill University’s Esophageal and Gastric Data- and Bio-Bank, we compared these imputation methods for 44 variables of the Functional Assessment of Cancer Therapy-Esophageal patient-reported outcome measure on execution time, distribution preservation, correlation maintenance, imputation accuracy, and clinical classification performance. Our comprehensive validation framework provides evidence-based recommendations for selecting appropriate imputation methods for esophageal cancer PRO research, which may improve the validity and reliability of research findings in this domain.

## 1. Introduction

Missing data is common when collecting patient-reported outcomes (PROs), which can not only effect statistical power but can also potentially bias results and our interpretation of them^1;2^. PROs are collected through standardized survey instruments (i.e., patient-reported outcome measures) with the intention of capturing patients’ self-reported symptom burden, functional status, and the impact that these have on their daily activities and quality-of-life^3^. Missing data occurs when values are recorded for some items or subjects but not for all items or subjects in a dataset^4^.

The type of missingness - namely, Missing Completely At Random (MCAR), Missing At Random (MAR), or Missing Not At Random (MNAR) - varies based on the underlying reason for non-response^5^. When missingness occurs due to factors such as technical difficulties or survey oversight, it is typically considered MCAR, meaning it is unrelated to the outcome of interest. In the case of PROs, for example, a patient might not receive a survey because the clinic was too busy. In contrast, MNAR is characterized by a strong association between missing data and the outcome of interest. Again, in the case of PROs, a patient may not receive a survey if they are too incapacitated to make a clinic visit.

In controlled clinical trials, missing PROs data can be mitigated through rigorous study design, protocol integration, and quality assurance, often aided by the strategic collection of auxiliary data to support assumptions about MAR mechanisms^6^. However, real-world clinical registries – i.e., the systematic collection of patients’ health and health care use, reflecting routine clinical practice – lack such stringent controls and dedicated resources. These registries face more complex and varied patterns of missingness driven by a wide range of potential causes and fewer dedicated resources for routine data checks.

Real-world clinical registries are more vulnerable to bias and reduced precision if missingness is ignored and statistical analyses only on subjects with complete data^7^. Consequently, robust post-hoc imputation strategies, such as multiple imputation (MI), and a detailed understanding of non-response mechanisms are essential for valid statistical inference in real-world registry analyses^5^. Simple methods like complete case analysis can yield inaccurate or inconsistent conclusions depending on the extent of missing data.

MI is a widely adopted statistical method for addressing missing data that involves imputing multiple plausible values for missing observations^4;8^. This process creates multiple complete datasets, each with the same size as the original sample, where all missing data have been replaced with plausible values. Statistical analyses are then conducted on each complete dataset, and the results are pooled across samples.

MI generally assumes MAR conditions and linear relationships among variables^9;10^. Under these MAR assumptions, PRO data present additional complexities including ordinal scales with discrete, bounded responses; potential ceiling or floor effects; and complex non-linear relationships between symptom domains^11;12^. Recent advances in machine learning offer promising new approaches to impute missing PRO data because they better preserve the complex relationships in PRO data through their ability to model non-linear associations, handle non-parametric distributions, and capture intricate patterns in high-dimensional sparse data^13^.

The primary objective of this study is to compare traditional statistical methods and more recent machine learning methods for imputing missing real-world PRO data. Our comprehensive benchmark evaluation extends beyond simple imputation accuracy to include computational efficiency, distribution preservation, correlation maintenance, and clinical classification performance. This will provide a holistic assessment of method suitability for different research contexts. The results from this study will support clinical researchers faced with missing PRO data make an informed decision based on their specific research needs, computational constraints, and analytical priorities.

## 2. Literature Review

Missing PRO data is pervasive in oncology trials, with 74.6% of breast cancer randomized controlled trials reporting missing data, yet only 19.6% explicitly documenting their statistical approach. This reveals a problematic preference for single imputation over more robust multiple imputation methods^14^. Extensive missing PRO data undermines trial validity by reducing statistical power to detect true treatment effects, increasing type II error rates, introducing systematic bias, and potentially compromising randomization^15^. These problems are amplified in cancer populations where disease progression, treatment toxicity, and mortality contribute to systematic patterns of missingness^16^, and retrospective data capture is often impossible^17^.

Multiple imputation (MI) has become the standard statistical approach for missing data in clinical research^18^, and is increasingly applied to missing PRO data in cancer trials^19^. However, implementation details significantly impact performance. For multi-item PRO instruments, researchers must decide whether to impute at the item level, subscale level, or composite score level, with optimal strategy depending on missingness patterns, sample size, and instrument structure^20^. For ordinal PRO data specifically, simple imputation methods such as mean substitution have proven inadequate, artificially improving psychometric properties and introducing bias^21–23^.

Real-world clinical registries face greater challenges than controlled trials due to less rigorous data collection protocols, more varied missingness patterns, and limited resources for data quality control^6^. Practical guidance for selecting appropriate imputation methods in these settings remains limited, with most comparative studies examining only narrow ranges of approaches. These studies usually compare variants of MI rather than systematically evaluating methods from diverse statistical and machine learning paradigms^19^. Despite extensive development of statistical methodology for missing PRO data, systematic comparisons across diverse paradigms remain scarce. Such comprehensive evaluations spanning traditional statistical methods, matrix factorization approaches, and modern deep learning architectures are still needed.

Machine learning approaches have shown promise for handling missing data in electronic health records^24^, yet their application to PRO-specific characteristics remains largely unexplored. These characteristics include ordinal scales with bounded responses, ceiling and floor effects, and theoretically-driven correlation structures across quality-of-life domains. The present study addresses this gap by providing the first comprehensive comparison of traditional statistical methods and modern machine learning approaches specifically for PRO data in a real-world cancer registry setting, evaluating performance across multiple clinically-relevant dimensions including comprehensive simulation and bootstrapping studies, computational efficiency, distribution preservation, correlation maintenance, imputation accuracy, and clinical classification performance.

## 3. Data

This study used data obtained from the McGill University’s Division of Thoracic and Upper Gastrointestinal Surgery (i.e., the Division), one of North America’s most comprehensive centres for esophageal cancer with over 250 new esophagogastric cancer referrals and 100 esophagectomies performed annually. Since 2006, the Division has maintained the Esophageal and Gastric Data- and Bio-Bank (EGDB Bank), a clinical database that includes longitudinal data collected from patients diagnosed with, and treated for, esophageal cancer. Patients provide informed consent for their data to be collected and used in future research studies. Nearly 100% of newly referred patients are approached for enrolment. This study protocol was approved by the McGill University Health Centre’s Centre for Applied Ethics.

### 3.1. Data Sources

Relevant to this study, the Division asks patients to complete the Functional Assessment of Cancer Therapy–Esophageal (FACT-E), a patient-reported outcome measure designed to assess symptom severity and impact in esophageal cancer patients, with established validity and reliability^25^. The FACT-E is comprised of four generic cancer domains (i.e., physical, social/family, emotional, and functional well-being) and one domain focused on symptoms specific to esophageal cancer. There are 44 items, each asking the respondent to rate their level of symptom severity over the past 7 days using a 5-point Likert scale. Responses to the items’ Likert scales are scored and aggregated according to the scoring guidelines^25–28^.

The Division collects the FACT-E at 11 different time points during the patient’s episode of care. Our analysis focused exclusively on one time point: “baseline” (i.e., collected at the patient’s first assessment with the thoracic surgeon). Baseline represents the most universal time point that every patient undergoes, providing the largest available sample size and avoiding temporal constraints that could bias certain methods. This cross-sectional focus ensures that specialized longitudinal methods are evaluated on their core imputation capabilities rather than their temporal modeling advantages.

### 3.2. Variables and Preprocessing

The dataset includes numerical and ordinal variables (Table 1). The numerical/ordinal variables (float or int type) included: id (i.e., an anonymous identifier given to each patient) and 44 FACT-E target variables comprising general physical well-being (i.e., variables = GP1-GP7), social/family well-being (i.e., variables = GS1-GS7), emotional well-being (i.e., variables = GE1-GE6), functional well-being (i.e., variables = GF1-GF7), and esophageal cancer-specific subscales (i.e., variables = A HN1-A HN5, A HN7, A HN10, A E1-A E7, A C6, A C2, A ACT11)^1^. Date variable qol date (i.e., the date that the FACT-E was completed) was retained and transformed into numerical features for the Da Xu et al.^29;30^ deep learning model, which incorporates date-derived features (year, month, day) as part of its architectural design.

**Table 1.** Percentage of missing values for each FACT-E variable.

| **GP1** | **GP2** | **GP3** | **GP4** | **GP5** | **GP6** | **GP7** | **GS1** | **GS2** | **GS3** | **GS4** |
| --- | --- | --- | --- | --- | --- | --- | --- | --- | --- | --- |
| 1.38 | 1.47 | 3.63 | 1.77 | 17.58 | 3.05 | 3.34 | 3.24 | 2.26 | 3.34 | 8.06 |
| **GS5** | **GS6** | **GS7** | **GE1** | **GE2** | **GE3** | **GE4** | **GE5** | **GE6** | **GF1** | **GF2** |
| 6.68 | 10.81 | 43.12 | 1.67 | 4.62 | 8.45 | 3.05 | 3.24 | 2.85 | 4.32 | 7.56 |
| **GF3** | **GF4** | **GF5** | **GF6** | **GF7** | **A_HN1** | **A_HN2** | **A_HN3** | **A_HN4** | **A_HN5** | **A_HN7** |
| 2.36 | 5.60 | 2.26 | 3.63 | 2.75 | 4.22 | 5.60 | 6.78 | 6.68 | 6.09 | 5.99 |
| **A_HN10** | **A_E1** | **A_E2** | **A_E3** | **A_E4** | **A_E5** | **A_E6** | **A_E7** | **A_C6** | **A_C2** | **A_ACT11** |
| 5.60 | 3.14 | 3.05 | 2.65 | 3.14 | 3.63 | 4.52 | 2.95 | 3.05 | 3.14 | 2.75 |

## 4. Imputation Model Formulation

We compared Multiple Imputation by Chained Equations (MICE)^31^, Variational Autoencoder (VAE)^32^, Denoising Autoencoder (DAE)^33^, Bayesian Principal Component Analysis (BPCA)^34^, a specialized Deep Learning approach based on recent literature^29;30^, SoftImpute^35;36^, and K-Nearest Neighbors (KNN)^37^. These methods were selected to represent diverse imputation paradigms and address the specific challenges of PRO data. MICE and KNN serve as established baseline methods widely used in clinical research. VAE and DAE represent modern deep learning approaches for complex data reconstruction. BPCA addresses the high-dimensional nature of PRO instruments through dimensionality reduction. The specialized deep learning approach^29;30^, originally designed for longitudinal clinical data with patient-specific embeddings and temporal patterns, was included to test whether such longitudinal-based modeling techniques provide imputation benefits when applied to cross-sectional baseline data. SoftImpute was selected for its proven performance in matrix completion tasks with structured missing patterns, which commonly occur in PRO data due to systematic item non-response^35;36^. This combination ensures comprehensive coverage of statistical, machine learning, and deep learning paradigms for evaluating imputation performance on complex PRO data.

### 4.1. Model Configuration and Comparison Strategy

To ensure fair comparison across all imputation methods, we deliberately avoided extensive hyperparameter tuning and instead used standard, well-established default configurations for each approach. Our primary objective was not to optimize individual methods for marginal performance improvements, but rather to provide a comprehensive benchmarking comparison of popular and relatively recent missing data imputation techniques as they would typically be applied in clinical research settings. This approach reflects real-world usage scenarios where researchers seek reliable, generalizable methods without extensive method-specific optimization.

Specifically, for MICE, we used standard LightGBM parameters with 80 boosting rounds and a maximum depth of 10. VAE employed typical architecture dimensions (64→32→16→8) with standard learning rates (0.001) and batch sizes (64). DAE used conventional five-layer symmetric architectures with standard regularization. BPCA utilized adaptive component selection with established convergence criteria. The Da Xu et al. deep learning approach followed the original paper’s architectural specifications. SoftImpute employed adaptive rank selection with standard convergence thresholds. KNN used commonly recommended neighbor counts (n=5) with established distance metrics.

The following subsections detail the architectural design and implementation approach for each of the seven imputation methods evaluated (details in Supplementary Notes).

### 4.2. Computational Environment

All analyses were conducted on Narval Cluster in the Digital Research Alliance of Canada infrastructure using a single node with 5 CPU cores, 20GB RAM, and 1 GPU (NVIDIA A100-SXM4-40GB) with CUDA 12.2. Python 3.11.5 was used with relevant libraries including `scikit-learn`^38^, `PyTorch`^39^, and `miceforest`^40^.

### 4.3. MICE (Multiple Imputation by Chained Equations)

MICE is an iterative statistical method that creates multiple plausible datasets by modeling each incomplete variable as a function of all other variables through chained conditional distributions^31^. For variable *Y*_*j*_ with missing values, MICE fits: 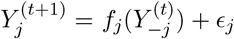, where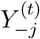 represents all other variables at iteration *t, f*_*j*_ is the prediction function, and *ϵ*_*j*_ is the error term.

We implemented MICE using the `miceforest` package^40^ with LightGBM^41^ as the base model for handling non-linear relationships. The implementation used 5 imputation iterations with all available variables as predictors. Hyperparameters were selected based on computational efficiency and model stability: 80 boosting rounds per model, maximum tree depth of 10, and 10 parallel threads for computation (OMP NUM THREADS = 10).

### 4.4. Variational Autoencoder (VAE)

VAE are generative deep learning models that learn a probabilistic latent representation of data through an encoder-decoder architecture with variational inference^32^. Unlike standard autoencoders, VAEs learn probabilistic distributions in the latent space, making them suitable for handling uncertainty in missing data imputation.

We implemented a VAE using `PyTorch`^39^ with threshold-based feature selection, excluding predictor columns with greater than 50% missingness while ensuring target columns were included regardless of missingness. All columns were converted to numeric format with initial mean imputation and z-score normalization. The encoder architecture follows systematic dimensionality reduction with hidden layers (64→32→16), learning increasingly abstract representations.

The encoder outputs two vectors: a mean vector (*μ*) and log-variance vector (log*σ*^2^), both of dimension 8, representing Gaussian distribution parameters in the latent space. The reparameterization trick computes *z* = *μ* + *σ* ⊙ *ϵ* where *ϵ* ∼ 𝒩 (0, 1). The decoder mirrors the encoder architecture in reverse (16→32→64→original dimension). We applied layer normalization, LeakyReLU activations^42^, and dropout (rate=0.1). The loss function combines masked mean squared error (MSE) reconstruction loss with KL divergence regularization *D*_*KL*_[*q*(*z* | *x*) ||*p*(*z*)]. Training used Adam optimizer (learning rate 0.001, maximum 100 epochs).

### 4.5. Denoising Autoencoder (DAE)

DAE are neural networks trained to reconstruct clean data from artificially corrupted inputs through a symmetric encoder-decoder architecture^33^.

We implemented a DAE using `scikit-learn`’s^38^ Multi-Layer Perceptron Regressor (MLPRegressor). We used RandomForestRegressor-based feature selection to identify up to 20 important predictors per target column, excluding columns with more than 30% missingness while ensuring target columns were included.

The implementation employed initial mean imputation with z-score normalization (StandardScaler). Selective noise addition was applied only to observed values: *X*_*noisy*_ = *X*_*scaled*_+ *ϵ* where *ϵ* ∼ 𝒩 (0, *σ*^2^) with noise factor *σ* = 0.1. The architecture features a five-layer symmetric structure (44→64→32→16→32→64→44) creating an hourglass shape with a bottleneck at 16 neurons (Fig. 1). We used ReLU activation functions. The loss function combines masked MSE reconstruction loss with L2 regularization: 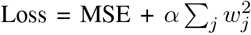, where *w*_*j*_ are the network weights and *α* = 0.0001 is the regularization strength to prevent over-fitting^33;43^. Training involved 200 iterations with Adam optimizer and automatic batch size determination (min(200, n samples)).

**Figure 1.**
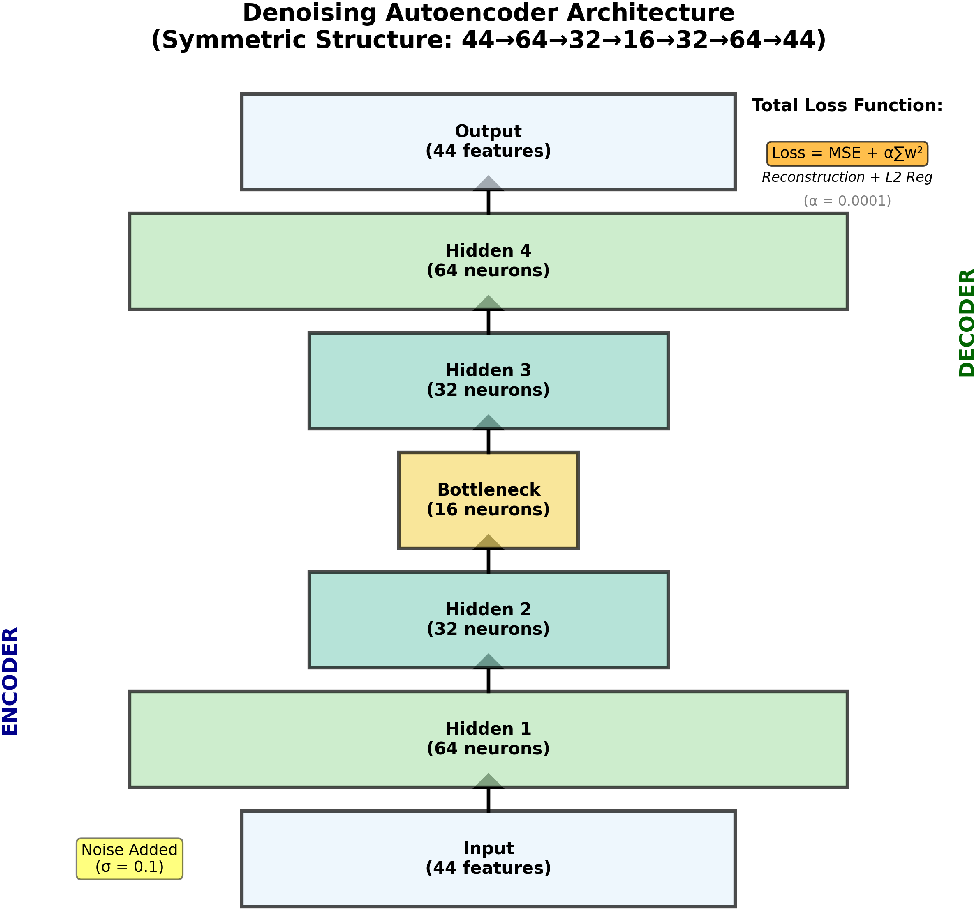
Denoising Autoencoder Architecture. The symmetric five-layer structure compresses input through encoder layers (44→64→32→16), reaches a bottleneck representation at 16 neurons, then reconstructs through decoder layers (16→32→64→44). Gaussian noise (*σ* = 0.1) is selectively added to observed values during training to improve generalization. The total loss function combines masked mean squared error (MSE) reconstruction loss with L2 regularization (*α* = 0.0001): 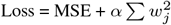, where *w*_*j*_ represents network weights.

### 4.6. Bayesian Principle Component Analysis (BPCA)

BPCA extends traditional PCA by treating model parameters as random variables with prior distributions, using variational inference to learn a probabilistic low-dimensional representation^34;44^.

We implemented BPCA using `PyTorch`^39^. Preprocessing involved initial mean imputation, z-score standardization, and standard PCA for parameter initialization. The model used all columns with less than 50% missingness, including all target columns. The probabilistic PCA formulation models observed data *X* ∈ ℝ^*n×d*^ as *X* = *ZW* ^*T*^ + *ϵ*, where *Z* ∈ ℝ^*n×k*^ are latent variables, *W* ∈ ℝ^*d×k*^ is the loading matrix, and *ϵ* represents observation noise.

The loading matrix *W* was modelled with variational parameters: mean *μ*_*W*_ ∈ ℝ^*d×k*^ and log-standard deviation log *σ*_*W*_ ∈ ℝ^*d×k*^. Noise precision *τ* = exp(log *τ*) was learned. Latent variables *z*_*i*_ ∼ 𝒩 (0, *I*) were initialized from standard PCA scores. We used *k* = min(5, *d* − 1) principal components.

Standard normal priors were used: *W*_*ij*_ ∼ 𝒩 (0, 1) for loadings and *z*_*i*_ ∼ 𝒩 (0, *I*) for latent variables. Posterior sampling drew 1,000 samples from 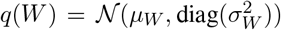. The reparameterization trick enabled gradient-based optimization by rewriting *W* ∼ *q*(*W*)

The loss function combined three components: ℒ = ℒ_recon_ + ℒ_*z*_ + ℒ_*W*_, where ℒ_recon_ = as *W* = *μ*_*W*_ + *σ*_*W*_ ⊙ *ϵ* where *ϵ* ∼ 𝒩 (0, *I*). *τ* Σ_*i,j∈*observed_(*X*_*ij*_ − [*ZW* ^*T*^]_*ij*_)^2^ is the masked reconstruction error weighted by noise precision, 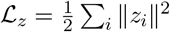 is the latent variable regularization, and 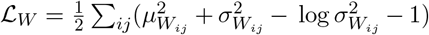 is the KL divergence between the approximate posterior and prior for loadings. Training used Adam optimizer (lr=0.01), ReduceLROnPlateau scheduler, early stopping (60 epoch patience), batch size min(64, n), and dynamic epochs max(100, min(300, 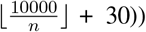

### 4.7. Patient-specific and temporal-patterned Deep Learning Autoencoder

Based on Da Xu et al.^29;30^, we implemented an autoencoder architecture combining patient-specific embeddings with temporal pattern modeling, originally designed for longitudinal clinical data but applied here to cross-sectional baseline data. Our implementation used columns with less than 50% missingness plus all target columns plus patient ID and qol date. Patient ID enables the model to learn patient-specific representations, while qol date provides temporal context (seasonal effects or cohort differences) that may influence PRO data reporting patterns even in cross-sectional data.

The architecture implements a patient-embedding autoencoder where input data *X* ∈ ℝ^*n×d*^ is processed alongside patient identifiers *P* ∈ {1, 2, …, *k*}^*n*^ where *k* = 936 represents unique patients. Patient IDs are mapped to dense embeddings *E*_*p*_ ∈ ℝ^16^ through an embedding layer. Temporal features (year, month, day) are extracted from qol date. The autoencoder follows a symmetric five-layer structure (64→32→16→32→64) where the input is compressed through *h*^(*l*+1)^ = *f* (*W* ^(*l*)^*h*^(*l*)^ + *b*^(*l*)^) with ELU activation function^45^. The decoder concatenates patient embeddings with latent code: 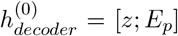 where *z* ∈ ℝ^16^. We employed BatchNorm, ELU activations, and dropout (rate=0.3).

The loss function combines two components: ℒ = ℒ_*recon*_ + *λ*ℒ_*temporal*_, where 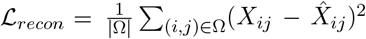 is the masked reconstruction loss on observed values Ω, and 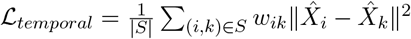 is the temporal similarity loss, where *S* represents patient pairs, *w*_*ik*_ are similarity weights based on patient proximity and temporal closeness, with *λ* = 0.4 and L2 weight decay (*α* = 0.0001).

Training used Adam optimizer (learning rate = 0.001), ReduceLROnPlateau scheduler, early stopping (20-epoch patience), 100 epochs, batch size 16, and 80/20 train/validation split.

### 4.8. SoftImpute

SoftImpute is a matrix completion algorithm that iteratively applies soft-thresholded singular value decomposition (SVD) to find low-rank approximations^35;36^.

We utilized Travis Brady’s SoftImpute implementation^46^, which employs iterative soft-thresholded SVD^47^. The SVD decomposes matrix *X* ∈ ℝ^*n×m*^ as *X* = *UDV* ^*T*^, where *U* ∈ ℝ^*n×r*^ and *V* ∈ ℝ^*m×r*^ are orthogonal matrices containing left and right singular vectors, and *D* ∈ ℝ^*r×r*^ is a diagonal matrix of singular values *σ*_1_ ≥ *σ*_2_ ≥ … ≥ *σ*_*r*_ ≥ 0.

Our implementation utilized data-driven rank selection with *J* = max(2, ⌊min(*n*_*samples*_, *n*_*features*_)*/*4⌋) components. The algorithm follows these iterative steps: (1) Initialize missing values with column means and apply z-score standardization, (2) Compute SVD: 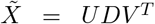, (3) Apply modified soft-thresholding: 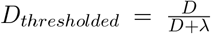 where *λ* = 0 for the base implementation, (4) Reconstruct matrix: 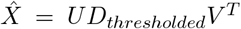, (5) Update missing values: 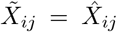, (6) Check convergence: ratio 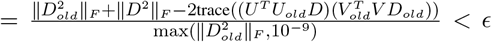 where *ϵ* = 10^*−*5^, or terminate after 100 iterations.

### 4.9. K-Nearest Neighbors (KNN)

KNN imputation is a non-parametric method that estimates missing values by identifying the k most similar complete observations based on available features and computing weighted averages^37^.

We implemented KNN imputation using `scikit-learn`’s^38^ KNNImputer. The method imputes missing values for observation *x*_*i*_ with missing feature *j* by finding k-nearest neighbors and computing weighted averages: 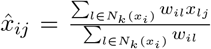, where *N*_*k*_(*x*_*i*_) represents the k-nearest neighbors of *x*_*i*_ and *w*_*il*_ are similarity weights.

Our implementation used *n*_*neighbors*_ = 5 with automatic adjustment based on data sparsity: *k*_*adjusted*_ = min(*k*, max(1, *C* − 1)), where *C* is the number of complete cases available for each variable. We used uniform weighting (*w*_*il*_ = 1) and nan euclidean distance metric to handle missing values in distance calculations: 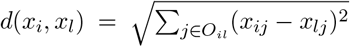, where *O*_*il*_ = {*j* : *x*_*ij*_ ≠ NA ∧ *x*_*lj*_ ≠ NA}.

## 5. Evaluation Framework

To comprehensively evaluate imputation performance, we implemented two complementary assessment approaches specifically designed for high-missingness data, incorporating both continuous accuracy measures and clinically-relevant classification metrics.

### 5.1. Direct Dataset Assessment

Our direct dataset evaluation used the original dataset’s naturally existing missingness patterns without cross-validation or artificial masking. We analyzed execution time for computational efficiency. Distribution similarity was evaluated using Kolmogorov-Smirnov (KS) tests^48^, comparing observed values against imputed values for each variable using the test statistic *D* = max_*x*_ |*F*_1_(*x*) − *F*_2_(*x*)| to measure the maximum difference between cumulative distribution functions *F*_1_ (observed values) and *F*_2_ (imputed values). Correlation preservation was analyzed by comparing correlation matrices before and after imputation, calculating absolute differences. Visualizations included histograms comparing imputed versus observed distributions and heatmaps showing correlation differences across methods.

### 5.2. Sparse Cross-Validation Approach

We developed a specialized cross-validation approach for datasets with moderate missingness rates, introducing artificial missingness by setting non-missing fold values to “NaN” for validation. Our approach performed column-wise validation, conducting separate 5-fold cross-validation for each of the 44 target variables. For each column, we identified all non-missing observations and randomly partitioned them into five folds, ensuring a minimum of 5 samples per fold or *n*_*observed*_*/*5 samples, whichever was larger. Columns with fewer than max(5, *n*_*folds*_ × 2) observations were excluded from validation.

For each imputation method, imputed values were compared against the original held-out values. We computed Mean Absolute Error (MAE) and Root Mean Square Error (RMSE) for each fold and method, along with execution time measurements. Results were averaged across all folds and columns.

### 5.3. Clinical Classification Metrics

Recognizing that FACT-E variables are ordinal scales (0-4), we implemented comprehensive classification evaluation by converting continuous imputed values to discrete clinical categories through rounding to the nearest integer and clipping to the valid range.

For each imputation method and variable, we calculated multiclass classification metrics including overall accuracy, multiclass Area Under the Curve (AUC) using a one-vs-rest approach, macro-averaged precision and recall. We computed sensitivity (true positive rate), specificity (true negative rate), positive predictive value (PPV), and negative predictive value (NPV) for each ordinal category, then averaged across categories.

The evaluation framework generated: quantitative accuracy measures (MAE, RMSE), computational efficiency (execution time), distribution preservation (KS test), correlation preservation, and classification performance (accuracy, AUC, sensitivity, specificity, PPV, NPV). A ranking system was implemented based on averaged ranks across all metrics, with lower ranks indicating better performance.

## 6. Simulation Study

We conducted two complementary simulation studies with 10 CPU cores, 40GB RAM, and NVIDIA A100 GPU. MICE threading was reduced to 2 threads per instance to enable 5 parallel jobs simultaneously, optimizing computational efficiency across available CPUs.

### 6.1. General Setup

We assessed imputation method stability using bootstrap resampling with 1,000 iterations. For each iteration, we resampled with replacement from the original dataset, preserving natural missingness patterns, then randomly masked 20% of observed values independently for each of the 44 FACT-E variables. All seven methods were applied to each bootstrap sample, and imputed values were compared against masked true values to calculate MAE, RMSE, and Kolmogorov-Smirnov (KS) statistics. We computed 95% confidence intervals using the percentile method and assessed stability using coefficient of variation (CV = SD/Mean).

For the second simulation, we extracted patients with complete FACT-E data for ground truth validation. For each of 1,000 simulation iterations, we artificially induced MCAR missingness by randomly deleting values at variable-specific rates matching the original dataset (1.38%-43.12%). All seven methods were applied, and imputed values were compared to known true values. We calculated bias (mean difference) and RMSE to quantify systematic error and prediction accuracy.

## 7. Results

The analytic dataset comprised 1,018 observations with 46 variables after preprocessing, representing data from 936 unique patients. Multiple baseline entries for some patients occurred due to clinical workflow variations (i.e., repeated assessments during pre-treatment evaluations or administrative scheduling), and all entries were retained given the small number of affected patients (n=73) and to preserve the natural data structure for realistic imputation evaluation. A total of 818 patients were excluded because they had no FACT-E data.

The target variables representing the complete FACT-E instrument (GP1-GP7, GS1-GS7, GE1-GE6, GF1-GF7, A HN1-A HN5, A HN7, A HN10, A E1-A E7, A C6, A C2, A ACT11) exhibited moderate missingness at the row level, with individual columns ranging from 1.38% (GP1) to 43.12% (GS7)^2^ missing values (see Table 1). Each variable had between 579-1,004 non-missing observations out of 1,018 total rows across the ordinal scale range 0-4, with low uniqueness ratios (0.5-0.9% unique values) characteristic of ordinal clinical scales.

### 7.1. Computational Performance

Execution times varied dramatically across methods (Fig. 2), reflecting fundamental differences in algorithmic complexity, methodological sophistication, and computational efficiency. KNN exhibited *O*(*n*^2^ × *m*) complexity (0.14 seconds) for pairwise distance calculations across sparse data, with *n* and *m* being data-derived parameters (i.e., *n* is the number of records and *m* is the number of features per record). The deep learning approaches, VAE and DAE, have a computational complexity of *O*(*n* × *d* × *h* × *e*), which is typical of neural network training. The method proposed by Da Xu et al. exhibits a higher complexity of *O*(*n* × *d*^2^ × *h* × *e*), where *n* and *d* are data-derived (sample size and input dimension), while *h* (hidden layer size) and *e* (epoch count) are user-tuned hyperparameters. DAE required 0.68 seconds with a scikit-learn implementation, while the Da Xu et al. method required 15.48 seconds due to additional computational burden from patient-specific embedding learning and temporal similarity calculations. VAE completed training at 8.49 seconds, benefiting from GPU acceleration on the NVIDIA A100-SXM4-40GB hardware and efficient batch processing where available.

**Figure 2.**
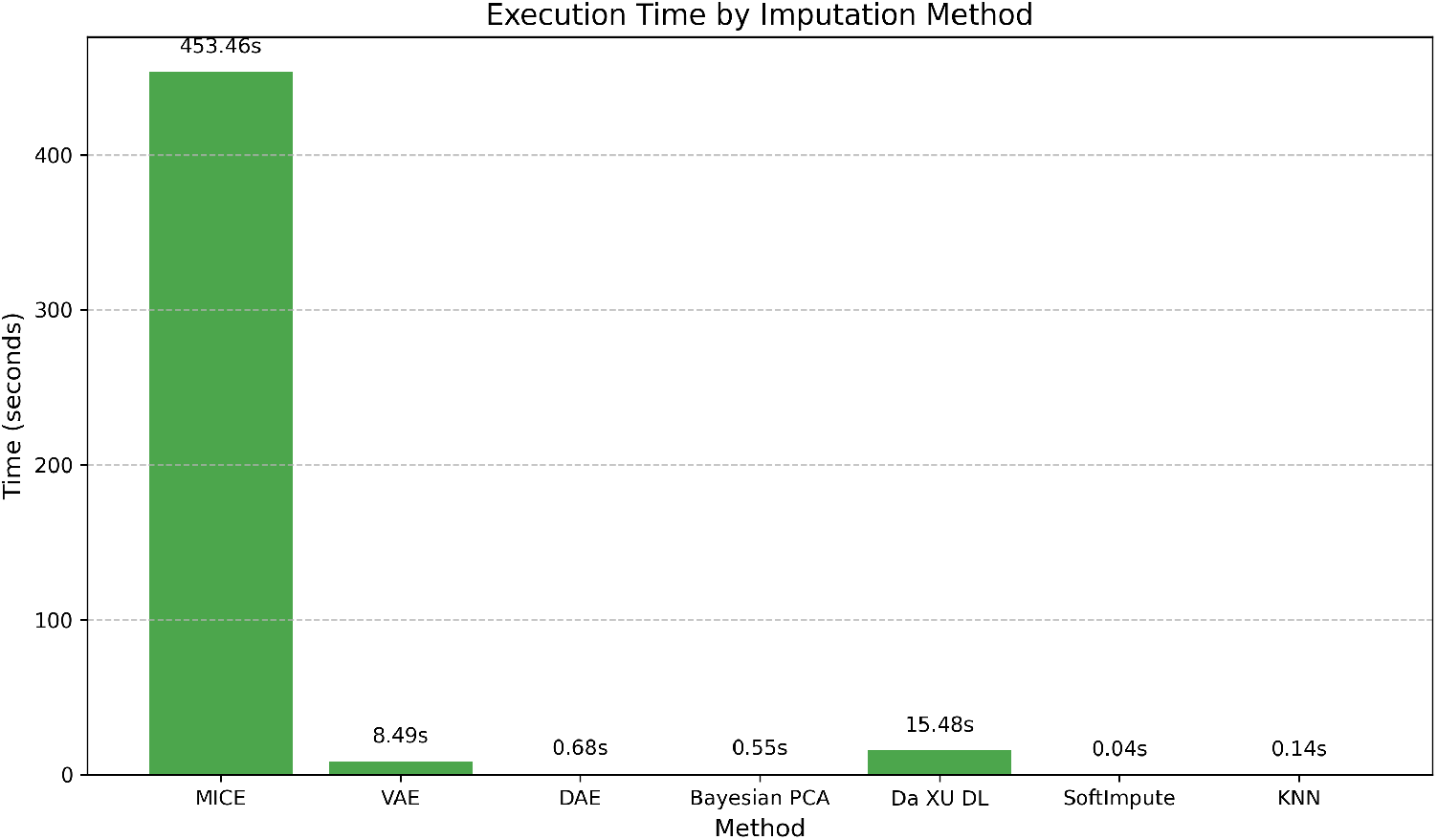
Execution Time by Imputation Method. Computational efficiency comparison across seven imputation methods applied to FACT-E quality-of-life data with moderate missingness rates (1.38%-43.12%). MICE required the longest execution time (453.46s), while SoftImpute was fastest (0.04s). The substantial time differences highlight trade-offs between computational efficiency and methodological complexity for clinical research applications.

MICE required the greatest amount of time - 453.46 seconds - due to its *O*(*k* × *m*^3^ × *n* log *n*) complexity, where *k* (iterations) and *m* (variables) are method parameters, *n* is data-derived sample size, and the log *n* term arises from multiple tree construction operations per iteration in the ensemble-based LightGBM implementation requiring sorted data structures for optimal split finding.

By comparison, the matrix factorization approaches demonstrated superior efficiency. Soft-Impute required 0.04 seconds with *O*(*k* × min(*n*, × *m*)^2^ *r*) complexity, where *r* (rank) and *k* (iterations) are method parameters, followed by Bayesian PCA at 0.55 seconds with *O*(*k* × *n* × *d*) complexity due to its iterative gradient-based optimization over latent variables and loading matrices, where *k* is the number of training iterations.

### 7.2. Distribution Preservation

The KS test (Fig. 3) results revealed substantial differences in distribution preservation capabilities across methods. MICE demonstrated superior performance with the lowest KS statistics across most variables (typically ranging from 0.05-0.35), indicating excellent preservation of the original data characteristics. Many MICE comparisons yielded p-values above 0.05 (Fig. S1), meaning the null hypothesis of identical distributions could not be rejected for numerous variables, demonstrating that imputed values closely matched the observed data distribution.

**Figure 3.**
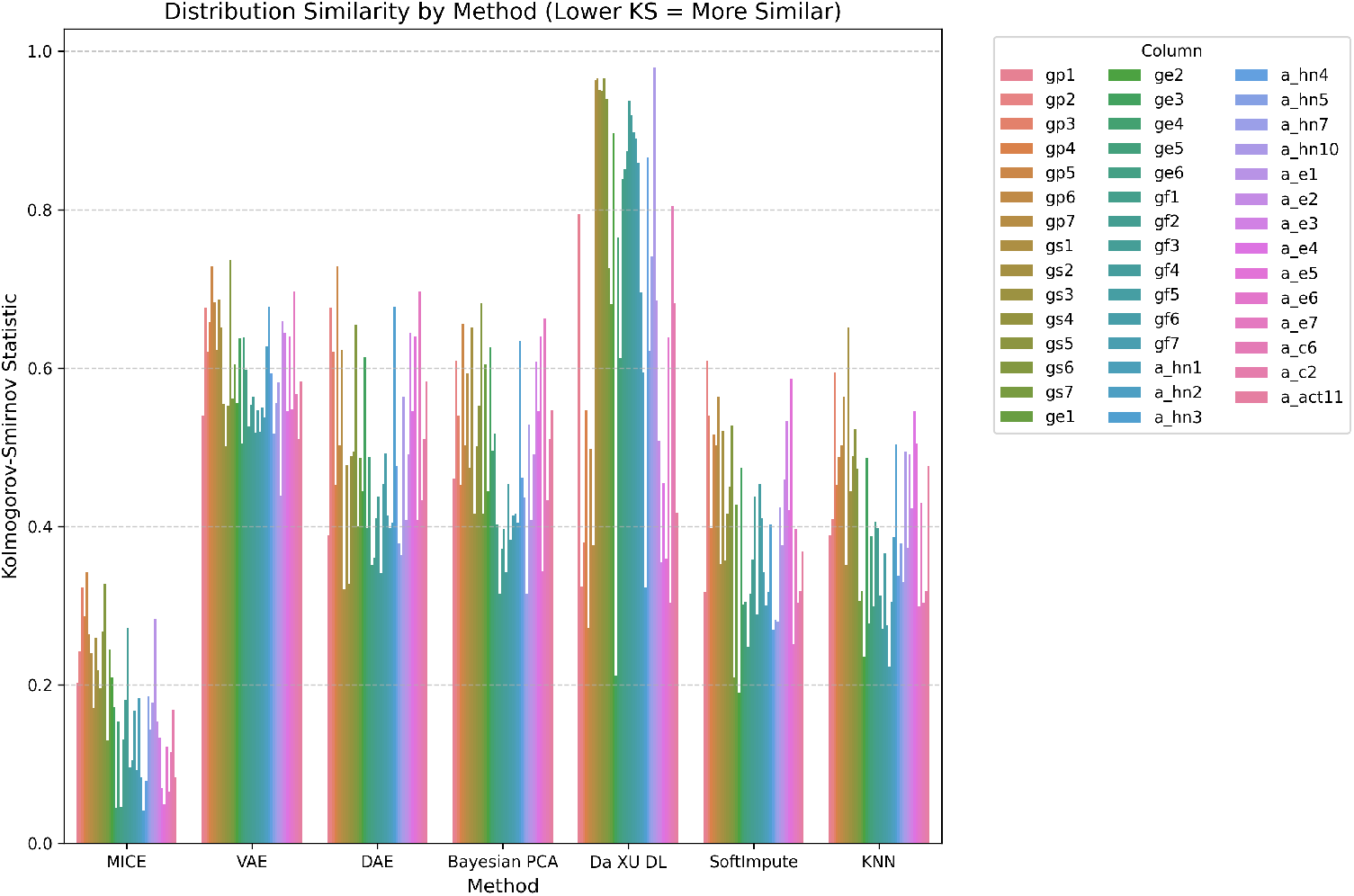
Distribution Similarity by Imputation Method. Kolmogorov-Smirnov test statistics comparing observed versus imputed value distributions across 44 FACT-E variables (lower values indicate better distribution preservation). MICE demon-strated superior performance with consistently low KS statistics (mostly below 0.35), while Da Xu et al. method^29;30^ showed poor preservation with values frequently reaching 0.85-0.95. VAE, DAE, and Bayesian PCA exhibited moderate performance with most variables in the 0.35-0.7 range, while SoftImpute and KNN showed intermediate to good performance with most variables below 0.6. Results highlight substantial differences in methods’ ability to maintain original data distributions.

VAE, DAE, and Bayesian PCA exhibited moderately high KS statistics (typically 0.35-0.7 range) with p-values *<* 0.1 across virtually all variables, indicating significant distributional differences from the observed data. These methods showed consistent patterns of moderate distribution preservation challenges across all FACT-E subscales, suggesting some difficulty with maintaining the discrete ordinal nature of the clinical scales.

The Da Xu et al. deep learning method showed the most extreme distribution preservation problems, with KS statistics frequently reaching 0.85-0.95 across many variables, particularly poor performance on social/family variables, functional and esophageal-specific items. This represents the worst distribution preservation among all evaluated methods, suggesting severe overfitting or inadequate model architecture for these clinical domains.

SoftImpute and KNN demonstrated intermediate to good performance with more consistent results across different variable types. SoftImpute showed KS statistics generally in the 0.2-0.6 range with relatively consistent performance, while KNN demonstrated similar performance (typically 0.25-0.6 range) with less variability across different clinical scales, suggesting both methods provide reasonably robust performance across diverse FACT-E domains.

### 7.3. Cross-Validation Performance

The sparse cross-validation framework successfully evaluated imputation accuracy across varying missingness patterns despite the computational intensity of validating 44 variables with 5-fold cross-validation across seven methods. The results revealed consistent performance patterns across methods (Fig. 4).

**Figure 4.**
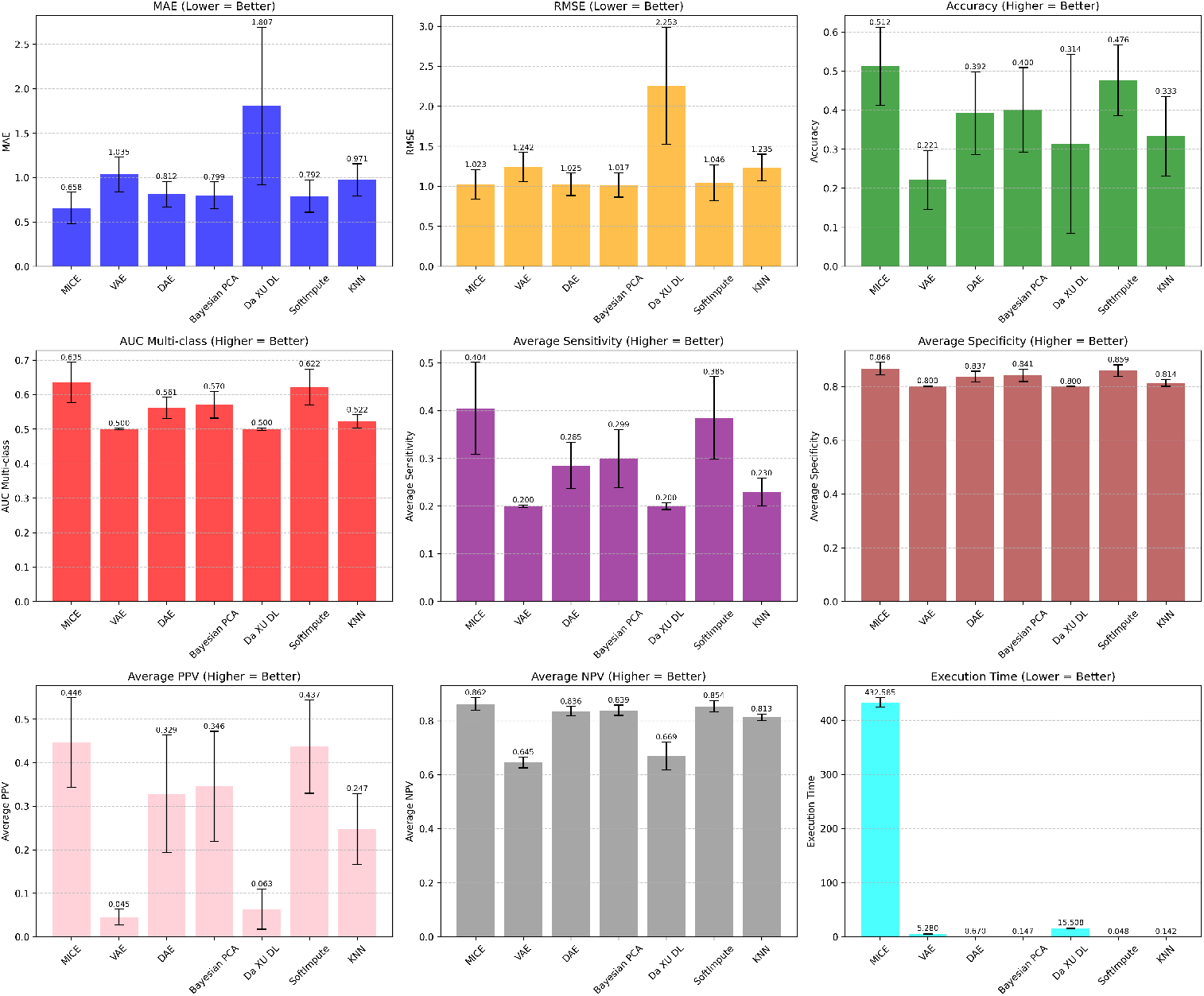
Comprehensive Performance Evaluation Across Seven Imputation Methods. Nine performance metrics from sparse cross-validation on 44 FACT-E variables with 1.38-43.12% missingness. MICE achieved the best performance with lowest MAE (0.658), RMSE (1.023), and highest accuracy (0.512) and AUC (0.635). SoftImpute showed competitive performance (accuracy: 0.476) with exceptional efficiency (0.048s). DAE, Bayesian PCA, and KNN demonstrated moderate performance, while Da Xu et al. method consistently exhibited poor accuracy (MAE: 1.807) and classification performance (accuracy: 0.314) despite high computational cost (15.5s). VAE showed inconsistent performance - showing satisfactory results in MAE and RMSE, but not the others. Error bars represent cross-validation standard deviations.

MICE achieved the most stable performance with a MAE of 0.658 and a RMSE of 1.023, demonstrating robust accuracy regardless of the specific clinical domain or missingness pattern. VAE, DAE, Bayesian PCA, SoftImpute, and KNN showed similar moderate performance with MAE values of 1.035, 0.812, 0.799, 0.792, and 0.971, respectively; and RMSE values of 1.242, 1.025, 1.017, 1.046, and 1.235. DAE, Bayesian PCA and SoftImpute demonstrated competitive accuracy, the lowest MAE and RMSE among non-MICE methods, indicating that despite its computational efficiency, it maintains reasonable imputation quality.

The Da Xu et al. deep learning method exhibited the poorest continuous accuracy metrics with MAE of 1.807 and RMSE of 2.253, showing highly variable performance consistent with the distribution preservation findings. This poor performance can be attributed to several methodological factors: the patient embedding layer may have suffered from overfitting due to sparse patient representation, the temporal similarity loss component potentially introduced noise rather than meaningful signal in this cross-sectional study with single time-point data, and the complex architecture with patient-specific parameters may have been inadequately suited for the extreme sparsity patterns in FACT-E data where the model struggled to learn meaningful patient-specific representations from limited observed values.

### 7.4. Clinical Classification Performance

MICE demonstrated superior classification performance with an accuracy of 0.512, outperforming all other methods. The method achieved balanced sensitivity (0.404) and high specificity (0.866), indicating a strong ability to correctly identify positive and negative cases across the ordinal categories. The multiclass AUC of 0.635 further confirmed MICE’s superior discriminative ability. PPV was 0.446, while NPV was 0.862 (Fig. 4).

SoftImpute showed the second-best classification performance with an accuracy of 0.476 and a multiclass AUC of 0.622. DAE and Bayesian PCA showed similar moderate classification performance with accuracy values of 0.392 and 0.400, respectively; and AUC values of 0.561 and 0.570, respectively. While these two methods showed intermediate sensitivity (0.285-0.299), they maintained high specificity (0.837-0.841), indicating reliable identification of patients without specific symptoms. KNN showed moderate performance across most metrics as well, with accuracy of 0.333 and AUC of 0.522.

VAE demonstrated poor classification performance with accuracy of 0.221 and AUC of 0.500, with low sensitivity (0.200). Da Xu et al. method also showed poor performance with an accuracy of 0.314 and AUC of 0.500, with particularly highly variable unstable performance across many metrics, suggesting challenges in correctly identifying patients with specific symptom severities. Both methods show suboptimal PPV and NPV values.

### 7.5. Correlation Preservation Analysis

The correlation preservation analysis revealed significant differences in how well each method maintained the natural relationships between FACT-E variables (Fig. 5, 6).

**Figure 5.**
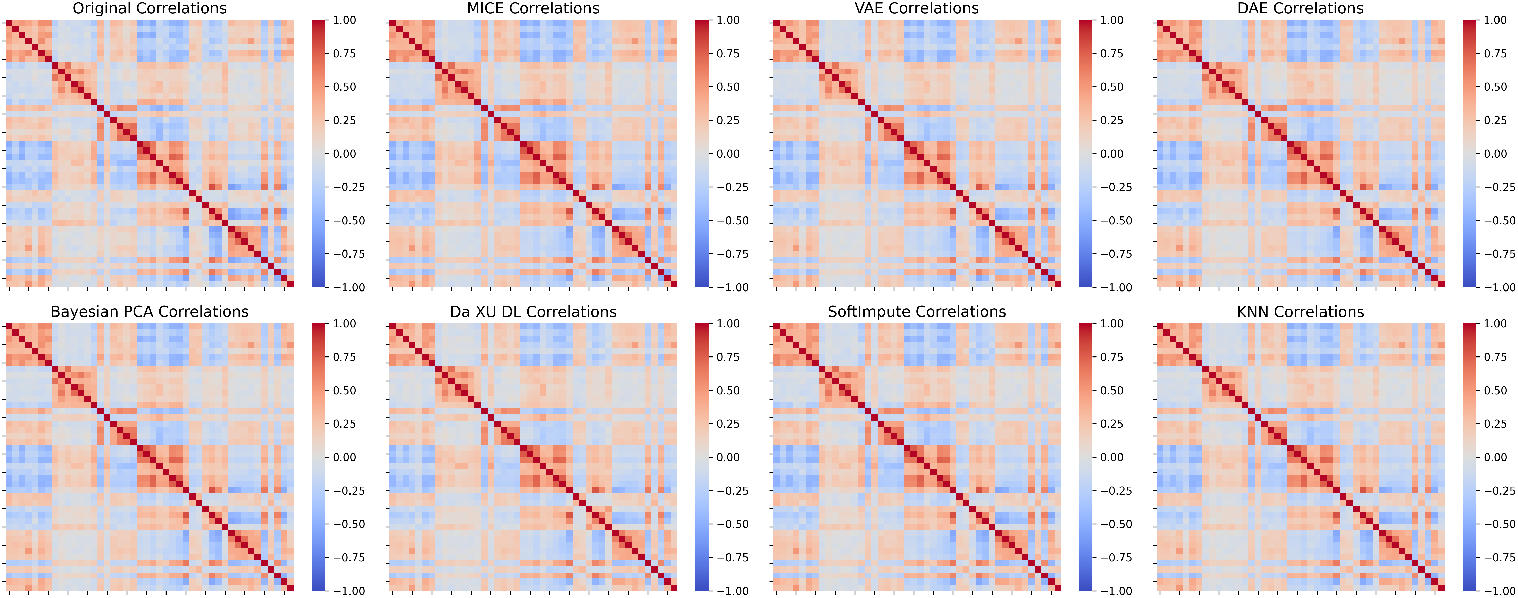
Correlation Matrix Across Imputed Results and Baseline Correlations in the Original Data. Heatmaps comparing correlation structures between 44 FACT-E variables for original data (baseline correlations observed in complete cases) and each imputation method. The variables are ordered as: GP1-GP7 (physical well-being), GS1-GS7 (social/family well-being), GE1-GE6 (emotional well-being), GF1-GF7 (functional well-being), A HN1-A HN5, A HN7, A HN10 (head/neck cancerspecific), A E1-A E7 (esophageal cancer-specific), A C6, A C2, A ACT11 (additional cancer concerns). Red indicates positive correlations, blue indicates negative correlations, with intensity representing correlation strength (-1 to +1). All methods maintained correlation patterns reasonably similar to the original structure in the less sparse original dataset with 936 patients.

**Figure 6.**
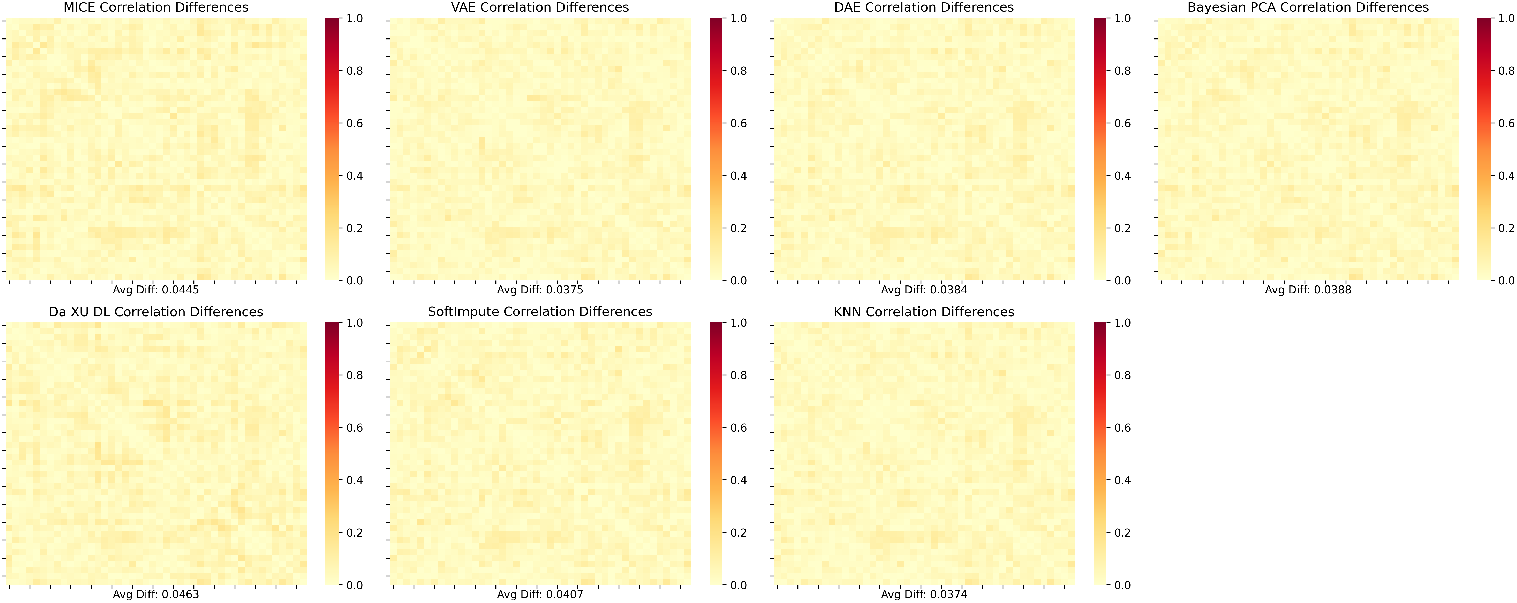
Absolute Correlation Differences Between Original Data and Each Imputed Matrix. Absolute differences between original and imputed correlation matrices (lower values indicate better preservation). Variable ordering is the same as Figure 5. VAE (0.0375), DAE (0.0384), Bayesian PCA (0.0388), and KNN (0.0374) achieved exceptional preservation with minimal deviation from original patterns. MICE (0.0445), and Da XU et al. method (0.0463), and SoftImpute (0.0407) showed good preservation. All methods demonstrated reasonable correlation preservation with relatively small differences, indicating that each approach maintains clinically meaningful relationships between FACT-E subscales in the less sparse original dataset with 936 patients.

All methods showed good correlation preservation, with average correlation difference between 0.0374 and 0.0463 (Fig. 5, 6). To assess robustness of each model under increased sparsity, a critical evaluation since real-world clinical datasets often exhibit varying and potentially higher missingness rates, we conducted an additional correlation preservation analysis using a more challenging sparsity-conditioned dataset with 1,836 observations from 1,754 unique patients by including previously excluded 818 patients, where missingness ranged from 45.3% to 68.5% across FACT-E variables (Fig. S2, S3).

VAE, DAE, Bayesian PCA, and KNN demonstrated superior correlation preservation with average differences of 0.0375, 0.0377, 0.0386, and 0.0382, respectively, maintaining correlation patterns nearly identical to the original data structure. SoftImpute showed moderate performance with an average difference of 0.0642, representing a slight degradation compared to the less sparse original dataset. The correlation matrices after these imputations closely resembled the baseline correlations observed in complete cases, indicating that these methods successfully preserved the established relationships between different FACT-E subscales without introducing artificial associations.

MICE exhibited sensitivity to increased sparsity, with correlation preservation degrading to an average difference of 0.1773. While the method maintained the general structure of correlations between variables, it exhibited notable deviation from the original correlation patterns, particularly in the strength of certain associations. MICE maintains general structure but deviates in specific associations as a consequence of its iterative approach under sparse FACT-E data conditions. However, the overall correlation framework remained clinically recognizable and interpretable.

The Da Xu et al. deep learning method exhibited the poorest correlation preservation among all evaluated methods, with an average correlation difference of 0.3334, showing concerning correlation distortions. The method introduced strong artificial correlations that were not present in the original data, particularly affecting relationships between different FACT-E domains. This pattern suggests potential overfitting to patient-specific patterns rather than learning generalizable quality-of-life structures, which could compromise the clinical validity of imputed values.

### 7.6. Ranked Comparison

Each method was assigned a rank (1=best, 7=worst) for each of the 12 performance metrics (MAE, RMSE, Accuracy, AUC, Sensitivity, Specificity, PPV, NPV, Time, KS Statistic, KS p-value, correlation differences) where lower ranks indicate better performance for MAE, RMSE, time, KS statistic, and correlation differences, while higher ranks indicate better performance for all other metrics. The arithmetic mean was calculated as: 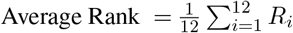, where *R*_*i*_ is the rank for metric *i*. Table 2 presents the ranking results across the complete evaluation framework.

**Table 2.** Comprehensive Summary of Imputation Method Performance Across All Evaluation Metrics.

| Method | MAE | RMSE | Accuracy | AUC | Sensitivity | Specificity | PPV | NPV | Time (s) | Avg KS Stat | Avg KS p-value | Sparse Correlation Differences | Avg Rank |
| --- | --- | --- | --- | --- | --- | --- | --- | --- | --- | --- | --- | --- | --- |
| MICE | 0.658 | 1.023 | 0.512 | 0.635 | 0.404 | 0.866 | 0.446 | 0.862 | 453.457 | 0.171 | 0.388 | 0.177 | 2 |
| SoftImpute | 0.792 | 1.046 | 0.476 | 0.622 | 0.385 | 0.859 | 0.437 | 0.854 | 0.044 | 0.389 | 0.006 | 0.064 | 2.417 |
| Bayesian PCA | 0.799 | 1.017 | 0.4 | 0.57 | 0.299 | 0.841 | 0.346 | 0.839 | 0.548 | 0.492 | 1.31e-4 | 0.039 | 3.333 |
| DAE | 0.812 | 1.025 | 0.392 | 0.561 | 0.285 | 0.837 | 0.329 | 0.836 | 0.679 | 0.488 | 6.35e-4 | 0.038 | 3.833 |
| KNN | 0.971 | 1.235 | 0.333 | 0.522 | 0.23 | 0.814 | 0.247 | 0.813 | 0.135 | 0.404 | 0.006 | 0.038 | 4.167 |
| VAE | 1.035 | 1.242 | 0.221 | 0.5 | 0.2 | 0.8 | 0.045 | 0.645 | 8.486 | 0.591 | 6.42e-06 | 0.038 | 6 |
| Da Xu DL | 1.807 | 2.253 | 0.314 | 0.5 | 0.2 | 0.8 | 0.063 | 0.669 | 15.479 | 0.68 | 0.002 | 0.333 | 6.25 |

*Note: Methods ranked by average rank across all metrics (lower is better). MAE = Mean Absolute Error; RMSE = Root Mean Square Error; AUC = Area Under Curve (multiclass), PPV = Positive Predictive Value, NPV = Negative Predictive Value, KS Stat = Kolmogorov-Smirnov statistic for distribution preservation (lower is better), KS p-value = significance level for distribution similarity test (higher is better). All continuous metrics are rounded to 3 decimal places.*

### 7.7. Bootstrap Stability Results

Bootstrap resampling confirmed robust method rankings across 1,000 iterations (Table S1). MICE maintained superior performance (MAE: 0.458, 95% CI: [0.430, 0.484]; RMSE: 0.837, 95% CI: [0.805, 0.867]) with low variability (CV *<* 0.03), followed by SoftImpute (MAE: 0.783) and Bayesian PCA (MAE: 0.806) (Fig. 7), mirroring original cross-validation rankings (Fig. 4). Bootstrap values had cleaner validation with 20% uniformly masked values and 80% available predictors, compared to natural complex missingness with reduced predictor availability. This cleaner context particularly benefited local neighborhood-based methods: MICE (MAE improved 30% from original 0.658) and KNN (MAE: 0.869, improved 11% from original 0.971) both showed enhanced performance, as their iterative and similarity-based approaches excel when more clean predictor information is available.

**Figure 7.**
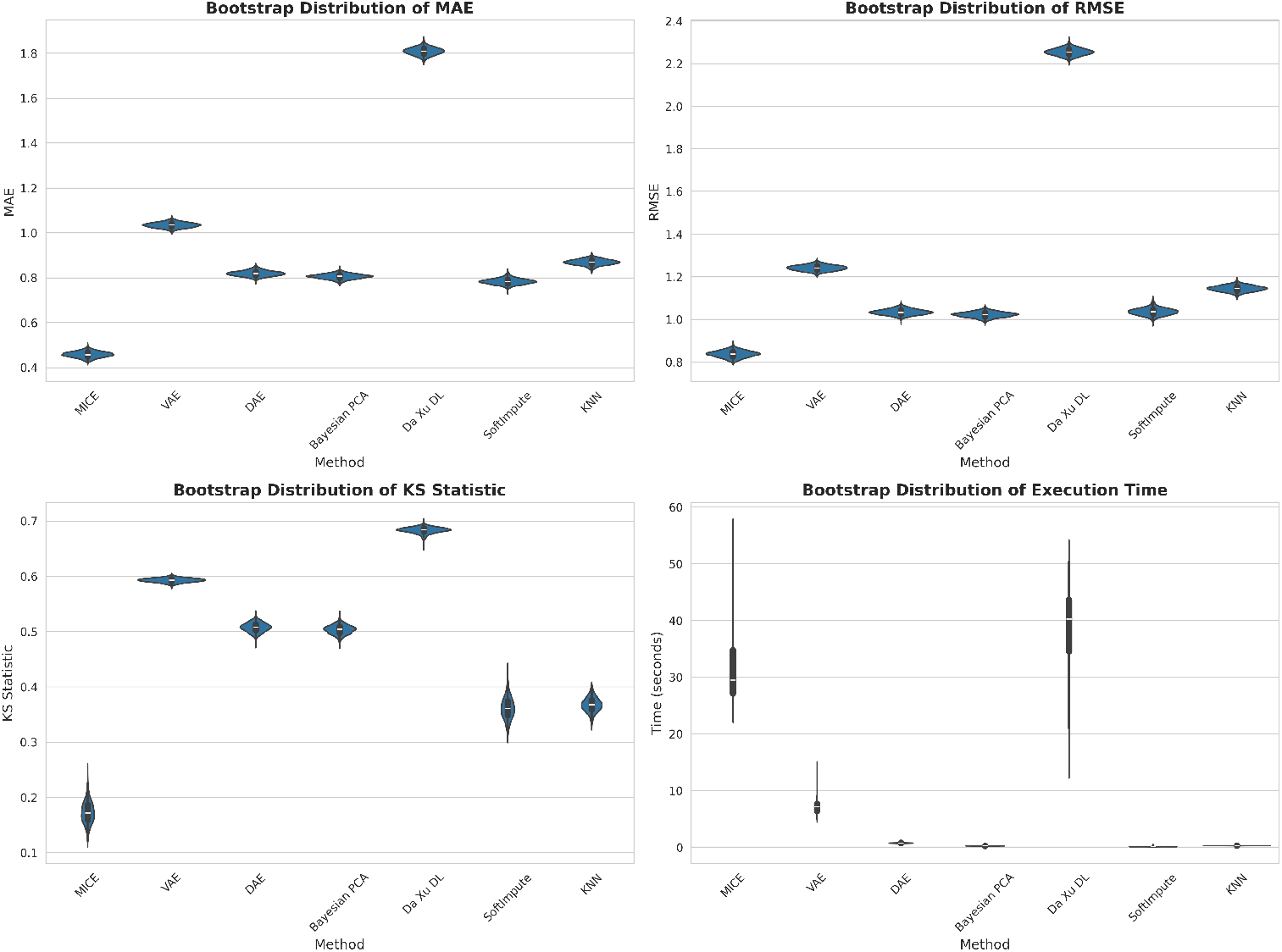
Bootstrap Assessment Across 1,000 Iterations. Distribution of performance metrics from bootstrap resampling with 20% uniform masking for validation. MICE achieved lowest MAE (0.458) and best distribution preservation (KS: 0.173), while Da Xu et al. method showed poorest performance (MAE: 1.809, KS: 0.684). Low coefficient of variation (CV *<* 0.03 for MAE/RMSE) across all methods confirms stable performance rankings. Da Xu et al. method exhibited high execution time variability (std 7.658s) due to patient embedding sensitivity to sample composition.

Distribution preservation patterns aligned with original findings (Fig. 3). MICE achieved best similarity (KS: 0.173, 95% CI: [0.138, 0.213]), Da Xu et al. deep learning showed poorest preservation (KS: 0.684, 95% CI: [0.670, 0.692]), and other methods demonstrated intermediate performance (KS: 0.361-0.593). Narrow confidence intervals confirmed consistent distribution preservation hierarchies across bootstrap samples.

Execution times differed notably from original analysis (Fig. 2) for MICE and Da Xu et al. deep learning. MICE required 32s mean (versus original 453s)—a 15-fold speedup from imputing only 20% masked values rather than full natural missingness, where MICE’s iterative approach benefits from cleaner missing patterns requiring fewer convergence iterations. Da Xu et al. deep learning showed high variability (mean: 38s, range: 13-54s, versus original 15s) due to bootstrap resampling creating unbalanced patient distributions: duplicated patients accelerate embedding convergence while absent patients prevent training, causing temporal similarity loss to require variable epochs (20-100), exposing the method’s brittleness to sample composition changes.

### 7.8. Complete-Case Simulation Results

Complete-case simulation (n=367, 1,000 iterations with MCAR-induced missingness matching original rates) revealed systematic bias patterns and method-specific sensitivities to data characteristics (Fig. 8 and Table S2). Most methods demonstrated near-zero bias: MICE (0.002), VAE (-0.002), DAE (-0.001), Bayesian PCA (0), SoftImpute (-0.001), and KNN (-0.005), with random errors canceling out across imputations. In contrast, Da Xu et al. deep learning exhibited severe systematic bias (-1.796), representing clinically unacceptable directional error of nearly 2 scale points on the 0-4 FACT-E range. This catastrophic failure stems from patient embedding overfitting, where the model memorizes patient-specific patterns that don’t generalize, causing consistent directional shifts in predictions rather than random errors. The temporal similarity loss, designed for longitudinal data, introduced additional noise in this cross-sectional context by pulling predictions toward artificially similar patients, further amplifying systematic bias.

**Figure 8.**
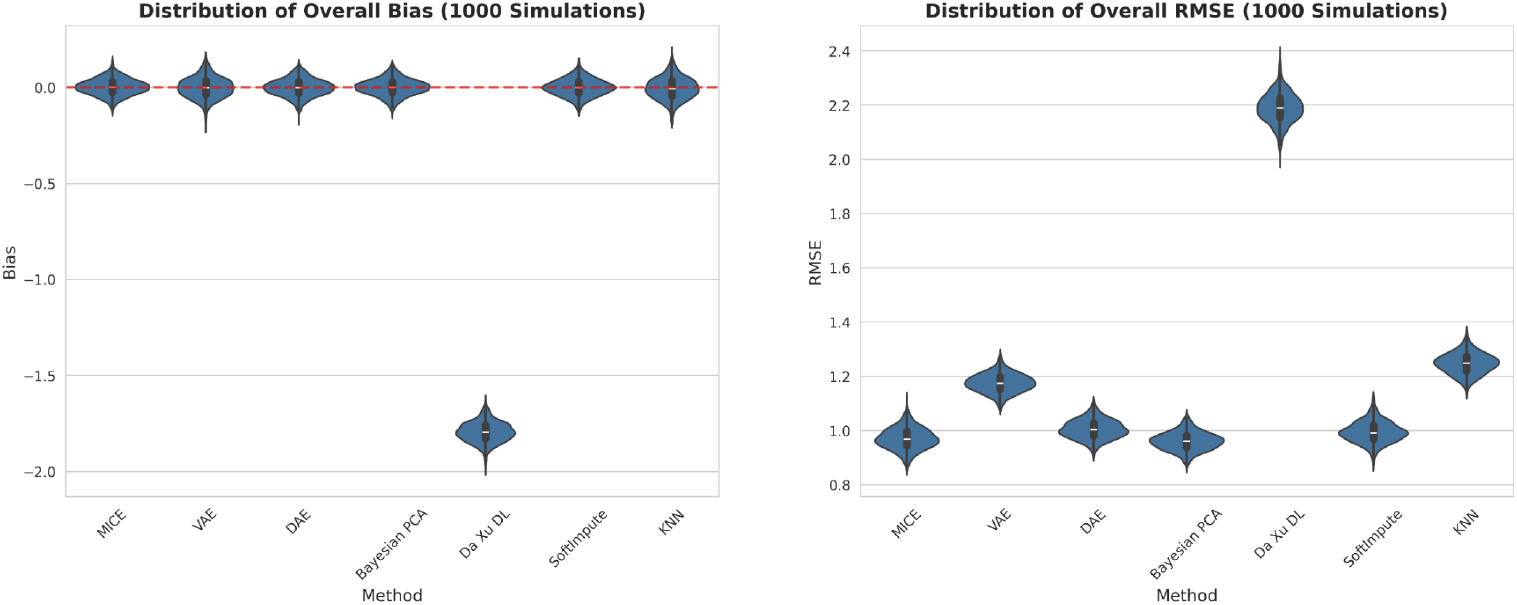
Complete-Case Simulation Across 1,000 Simulations. Imputation performance against known ground truth (n=367 complete cases, MCAR missingness matching original rates). Most methods showed near-zero bias (DAE: -0.001, Bayesian PCA: 0, SoftImpute: -0.001) and comparable RMSE (0.961-1.247), while Da Xu et al. method exhibited severe systematic bias (-1.796) and RMSE (2.189) due to patient embedding overfitting. Red dashed line indicates zero bias.

RMSE patterns mirrored bias findings: MICE (0.970), VAE (1.175), DAE (1.004), Bayesian PCA (0.961), SoftImpute (0.994), and KNN (1.247) showed comparable accuracy, while Da Xu DL (2.189) exhibited higher errors. Notably, MICE and KNN showed no performance improvement over bootstrap results, contrasting with other methods, though for different reasons. KNN, as a purely local similarity-based method, loses discriminative power in homogeneous complete-case populations where all patients are similar, reducing the advantage of finding “nearest neighbors.” MICE, while model-based rather than distance-based, relies on LightGBM’s tree-based ensemble learning that excels at capturing complex heterogeneous patterns and interactions; in the uniform complete-case setting with high item correlations and reduced variability, this complexity becomes unnecessary, offering no advantage over simpler approaches. Conversely, global latent structure methods (VAE, Bayesian PCA, SoftImpute) benefited from the strong, smooth low-rank covariance structure inherent to complete cases, with VAE slightly outranking KNN (reversed from Fig. 4 rankings).

## 8. Discussion

To the best of our knowledge, this study offers the first comprehensive evaluation of missing data imputation methods for PRO data in a real-world cancer registry. We evaluated seven approaches to impute missing data from the FACT-E, a common patient-reported outcome measure used for esophageal cancer. Our large registry had moderate missingness rates ranging from 1.38% to 43.12%. Rather than pursuing method-specific optimization for marginal performance gains, we used standard, generalizable configurations providing practical model selection guidance through performance baselines across 12 evaluation criteria, statistical significance testing for distribution preservation, and computational complexity analysis. The results demonstrate clear performance hierarchies across all methods and trade-offs between accuracy, efficiency, and clinical utility.

The results demonstrate that MICE emerged as the superior method across most evaluation criteria. This can be attributed to its iterative chained equations architecture leveraging LightGBM’s gradient boosting to capture complex non-linear relationships while maintaining distributional fidelity via ensemble averaging. MICE achieved exceptional distribution preservation and non-significant differences for multiple variables. This translates into superior clinical classification accuracy, sensitivity, and multiclass AUC, plus the lowest continuous error metrics, compared to the other methods tested. This makes it ideal for clinical applications needing accurate symptom severity categorization.

However, MICE showed sensitivity to increased sparsity due to iterative uncertainty propagation amplifying correlation distortions under sparse data conditions. In contrast, VAE, DAE, Bayesian PCA, and KNN maintained robust correlation preservation through their respective methodological strengths: VAE’s probabilistic latent space representation, DAE’s denoising regularization, Bayesian PCA’s probabilistic loading matrix, and KNN’s similarity-based local structure preservation. Additionally, MICE’s computational demands limit its use in largescale studies, being about 28 times slower than the Da Xu et al. method and over 9,800 times slower than SoftImpute. Matrix factorization methods like SoftImpute demonstrate superior computational scalability for large-scale clinical studies, while iterative ensemble methods face scalability challenges due to multiplicative complexity factors.

Deep learning methods showed mixed results. The complex Da XU et al. method exhibited instability with poor overall performance under increased sparsity conditions, while simpler VAE also underperformed compared to well-designed DAE. These findings indicate that deep learning methods for sparse clinical data require balanced architectural design. SoftImpute balanced efficiency and accuracy via low-rank approximation capturing global correlations, suiting large-scale clinical studies, while KNN provided stable, interpretable results through similarity-based imputation.

Based on the results from this study, we offer several recommendations: MICE for high-stakes applications prioritizing accuracy; SoftImpute for routine clinical use balancing accuracy and efficiency; and Bayesian PCA for research valuing correlation preservation and uncertainty quantification. The rankings highlights performance hierarchies to guide method selection by research priorities and constraints, serving as a template for future missing data research in PROs to enhance quality-of-life study validity and reliability in cancer care.

Despite comprehensive evaluation across seven imputation methods, the overall performance levels reveal fundamental challenges in real-world PRO data imputation. Even the best-performing method (MICE) achieved 51.2% classification accuracy, indicating that accurate imputation of PRO remains problematic across all evaluated approaches. The modest performance across all methods likely reflects the complexity of missing PRO data that extends beyond MAR assumptions. For instance, item GS7 of the FACT-E (“I am satisfied with my sex life”) exhibited disproportionate missingness (43.12%), suggesting MNAR patterns where patients may systematically avoid responding to sensitive questions. This is why the preservation of correlation structure is particularly important for PRO data, as the established relationships between physical, social, emotional, functional, and disease-specific subscales reflect well-documented clinical patterns in quality-of-life assessment. Such systematic missingness patterns indicate that traditional imputation approaches may be fundamentally limited for certain PRO domains. The results from this study suggest that considerably more research is needed into the statistical strategies for imputing missing real-world PRO data, including the implications for synthetic data simulation.

Several limitations should be considered when interpreting these results. Our analysis focused on baseline cross-sectional data from a single-center dataset, potentially limiting generalizability. The framework used artificial missingness under MAR assumptions, but real-world MNAR patterns could affect method performance. Using standard configurations ensured practical applicability but may not reflect optimal performance from extensive hyperparameter tuning. Given these limitations, researchers should exercise caution when applying any imputation method to PRO data, consider reporting results both with and without imputation, conduct sensitivity analyses to assess the impact of different imputation approaches on study conclusions, and acknowledge the inherent uncertainty in imputed values when making clinical inferences.

In conclusion, our comprehensive evaluation demonstrates that MICE emerges as the most robust imputation method for handling missing real-world PRO data in esophageal cancer, excelling in distribution preservation, imputation accuracy, and clinical classification performance. While machine learning approaches like DAE and SoftImpute offer viable alternatives for specific scenarios involving high-dimensional or sparse data, methods such as the Da XU deep autoencoder and VAE showed greater instability and resource demands, limiting their practical utility in routine clinical research. These findings provide evidence-based guidance for researchers to select appropriate imputation strategies tailored to their study constraints and priorities, ultimately enhancing the validity, reliability, and interpretability of PRO analyses in oncology. Future work should validate these methods in multi-center longitudinal datasets and under varying missingness mechanisms, including MNAR, to further refine recommendations and develop broader MI strategies for real-world PRO data.

## Authors’ contributions

Y.J.K. designed and implemented the methodology and analyzed the results. S.N., C.M., J.C.L., J.S., L.F., and M.D. collected the data. Y.S. and S.D. contributed equally to this work as second authors. E.A.M. and R.T.C. conceived and designed the study and supervised the production of results. Y.J.K., E.A.M., and R.T.C. were involved in writing and revising the manuscript and have approved the final version.

## 9. Data availability

The datasets analyzed in this study are not publicly available due to patient privacy and institutional data sharing restrictions. Requests for data access may be directed to the corresponding author and will be considered on a case-by-case basis following institutional review board approval and establishment of appropriate data use agreements.

## 10. Code availability

The source code and results reproduction are publicly available at our GitHub repository (https://github.com/Crump-Lab/missing_imputation).

## Acknowledgement(s)

Computations were made on the supercomputer Narval, managed by Calcul Québec and the Digital Research Alliance of Canada (Alliance). The operation of this supercomputer is funded by the Alliance, le ministère de l’Èconomie, de l’Innovation et de l’Ènergie du Québec (MEIE) and le Fonds de recherche du Québec (FRQ).

## Funding

This research was funded with generous support from the Rossy Cancer Network’s Cancer Quality and Innovation program.

## Ethics approval and consent to participate

This study was approved by the McGill University Health Centre’s Centre for Applied Ethics. All patients in the EGDB Bank provided informed consent for their data to be used in research studies.

## Appendices

### S1. MICE Implementation Details

#### Variable Schema

For each of the 44 FACT-E target variables, all other columns in the dataset were used as predictors through a variable schema configuration, ensuring the full correlation structure among FACT-E subscales was leveraged during imputation^31;49^.

#### Methodological Advantages

MICE provides theoretical guarantees for valid statistical inference when the missing at random assumption is satisfied and naturally handles different variable types within the same framework^50;51^. The method’s flexibility allows incorporation of auxiliary variables that may not be of direct interest but improve imputation quality. Its iterative nature enables capturing non-linear relationships through ensemble methods like LightGBM while maintaining computational efficiency through parallelization.

### S2. VAE Implementation Details

#### Reparameterization Trick

The reparameterization trick enables gradient-based training despite stochastic sampling by allowing gradients to flow through the sampling operation during backpropagation.

#### Architectural Rationale

Layer normalization was chosen for CPU efficiency, operating on individual samples rather than batch statistics. The *β*-VAE formulation for KL divergence encourages the latent space to follow a known distribution while maintaining sufficient representational capacity for disentanglement.

#### Methodological Advantages

VAE provides superior handling of non-linear relationships through its deep neural network architecture^32;52^. The latent space representation captures underlying patterns in PRO data including non-additive interactions between symptom domains, threshold effects, and clustering patterns. The KL divergence regularization prevents the model from generating unrealistic values that deviate significantly from observed data patterns^53^.

### S3. DAE Implementation Details

#### Feature Selection Strategy

For each target variable (e.g., GE1), missing values are imputed based on patterns learned from up to 20 selected columns, including target columns and other predictors with less than 30% missingness, identified using RandomForestRegressor. This differs from our VAE implementation, which includes all columns with less than 50% missingness.

#### Noise Injection Rationale

Selective noise addition to observed values serves as a denoising effect, improving generalization while preserving missing value patterns. The noise injection forces the model to learn more generalizable patterns, reducing overfitting on sparse data^54;55^.

#### Architectural Design

The symmetric hourglass architecture forces the model to learn compressed representations in the middle layer before reconstruction. Adaptive learning rate adjustment and automatic batch size determination balance computational efficiency with gradient stability.

#### Methodological Advantages

This approach provides simplicity, computational efficiency, and framework independence while learning robust representations through its denoising mechanism^33;56^. It is particularly practical for clinical research settings where computational resources may be limited, while still capturing non-linear relationships within FACT-E subscales^13^.

### S4. BPCA Implementation Details

#### Variational Framework

The loading matrix uses log-standard deviation to ensure positivity of variance parameters. The reparameterization trick enables gradient-based optimization by allowing gradients to flow through sampling operations during backpropagation.

#### Component Selection

The choice of *k* = min(5, *d* − 1) principal components captures main variation patterns while preventing overfitting in the FACT-E data structure.

#### Training Strategy

Dynamic epochs adapt computational effort to dataset size. Early stopping (60 epoch patience) prevents overfitting while ReduceLROnPlateau scheduler adjusts learning rate based on validation performance.

#### Methodological Advantages

BPCA provides principled uncertainty quantification through full posterior distributions for missing values, valuable for clinical decision-making. The method excels at capturing linear relationships and correlation structures in PRO data with strong inter-scale correlations^34;57^. Its computational speed makes it suitable for rapid prototyping and large-scale applications^44;58^.

### S5. Da Xu et al. Deep Learning Implementation Details

#### Patient Embedding

While 73 patients have multiple baseline entries in our dataset, all patient IDs are mapped to 16-dimensional embeddings to capture patient-specific characteristics.

#### Temporal Similarity Loss

The temporal loss calculates pairwise differences between predictions weighted by patient similarity (*w*_*ik*_ based on patient proximity and temporal closeness), encouraging similar imputations for patients with comparable characteristics at similar time points.

#### Cross-sectional Application

This method, originally designed for longitudinal clinical data, was applied to cross-sectional baseline data to test whether patient-specific modeling and temporal pattern recognition provide imputation benefits even without longitudinal follow-up. The approach evaluates whether techniques valuable for patient-specific disease progression trajectories can improve single time-point imputation quality.

### S6. SoftImpute Implementation Details

#### Rank Selection

The adaptive rank selection *J* = max(2, ⌊min(*n*_*samples*_, *n*_*features*_)*/*4⌋) ensures computational efficiency by limiting the number of factors while capturing essential data structure through the largest singular values that explain the majority of variance in the PRO data.

#### Modified Soft-thresholding

The modification 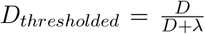 was chosen for improved numerical stability in our dataset, achieving shrinkage while preserving n-zero no gular vsainlu-es. This effectively performs nuclear norm regularization 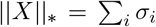, which encourages low-rank solutions by penalizing model complexity.

#### Convergence Criterion

The Frobenius norm ratio provides a robust convergence check based on the factorized matrices rather than the full imputed matrix, improving computational efficiency.

#### Methodological Advantages

SoftImpute excels in capturing global correlation patterns through matrix factorization, making it effective for FACT-E subscales. The method is theoretically grounded in convex optimization, providing guaranteed convergence and stable, reproducible results^35;59^. Its efficiency makes it well-suited for PRO datasets with high missingness rates, while the deterministic nature ensures reproducibility for clinical research applications.

### S7. KNN Implementation Details

#### Adaptive Neighbor Selection

The algorithm dynamically adjusts the number of neighbors using *k*_*final*_ = min(*k*_*original*_, max(1, min_*j∈J*_ (|{*i* : *x*_*ij*_ ≠ NA ∧ ∃*l* ≠ *j* : *x*_*il*_ ≠ NA}|−1))), where *J* represents the set of variables to be imputed, ensuring robust imputation even with sparse data.

#### Distance Metric

The nan euclidean distance metric computes distances using only dimensions where both observations have valid data, making it particularly suitable for sparse clinical datasets where different patients may have missing values in different variables. The automatic adjustment of *k* based on data availability ensures robustness even when few complete cases are available.

#### Methodological Advantages

KNN provides intuitive, non-parametric imputation based on patient similarity, making it interpretable for clinical applications. The method naturally adapts to local data structures and can capture complex, non-linear relationships without strong distributional assumptions^60;61^, making it robust to various data patterns in PROs. Its robustness to outliers and ability to work well with small sample sizes make it valuable for clinical datasets where extreme responses may be clinically meaningful. The non-parametric nature makes it suitable for ordinal clinical scales that may not follow normal distributions, and it can preserve local neighborhoods to capture patient subgroups with similar clinical characteristics.

---

**Table S1.** Bootstrap Stability Assessment Summary Statistics (1,000 Iterations)

| Method | Metrics | Mean | SD | 95% CI |
| --- | --- | --- | --- | --- |
| MICE | MAE | 0.458 | 0.013 | [0.430, 0.484] |
|  | RMSE | 0.837 | 0.016 | [0.805, 0.867] |
|  | KS Statistic | 0.173 | 0.019 | [0.138, 0.213] |
|  | Time (s) | 31.579 | 5.828 | [25.444, 45.784] |
| VAE | MAE | 1.035 | 0.012 | [1.011, 1.057] |
|  | RMSE | 1.241 | 0.013 | [1.217, 1.265] |
|  | KS Statistic | 0.593 | 0.004 | [0.585, 0.600] |
|  | Time (s) | 7.051 | 1.001 | [5.174, 8.690] |
| DAE | MAE | 0.819 | 0.012 | [0.797, 0.844] |
|  | RMSE | 1.034 | 0.013 | [1.009, 1.061] |
|  | KS Statistic | 0.508 | 0.008 | [0.491, 0.523] |
|  | Time (s) | 0.775 | 0.083 | [0.706, 0.975] |
| Bayesian PCA | MAE | 0.806 | 0.012 | [0.782, 0.828] |
|  | RMSE | 1.022 | 0.013 | [0.996, 1.046] |
|  | KS Statistic | 0.504 | 0.008 | [0.487, 0.518] |
|  | Time (s) | 0.229 | 0.028 | [0.190, 0.275] |
| Da Xu DL | MAE | 1.809 | 0.017 | [1.776, 1.842] |
|  | RMSE | 2.254 | 0.017 | [2.223, 2.287] |
|  | KS Statistic | 0.684 | 0.006 | [0.670, 0.692] |
|  | Time (s) | 38.103 | 7.658 | [18.235, 48.872] |
| SoftImpute | MAE | 0.783 | 0.013 | [0.758, 0.810] |
|  | RMSE | 1.036 | 0.017 | [1.005, 1.071] |
|  | KS Statistic | 0.361 | 0.019 | [0.326, 0.400] |
|  | Time (s) | 0.110 | 0.040 | [0.083, 0.228] |
| KNN | MAE | 0.869 | 0.013 | [0.843, 0.892] |
|  | RMSE | 1.144 | 0.014 | [1.118, 1.169] |
|  | KS Statistic | 0.368 | 0.013 | [0.342, 0.393] |
|  | Time (s) | 0.282 | 0.034 | [0.262, 0.407] |

*Note:* MAE = Mean Absolute Error, RMSE = Root Mean Square Error, KS = Kolmogorov-Smirnov statistic for distribution preservation (lower is better), Time = execution time per iteration. All values rounded to three decimal places. 95% CI calculated using percentile method.

**Table S2.** Complete-Case Simulation Summary Statistics (1,000 Simulations)

| **Method** | **Metrics** | **Mean** | **SD** | **95% CI** |
| --- | --- | --- | --- | --- |
| MICE | Bias | 0.002 | 0.042 | [-0.080, 0.081] |
|  | RMSE | 0.970 | 0.040 | [0.892, 1.054] |
|  | Time (s) | 19.397 | 4.098 | [15.438, 29.850] |
| VAE | Bias | -0.002 | 0.053 | [-0.100, 0.110] |
|  | RMSE | 1.175 | 0.034 | [1.106, 1.241] |
|  | Time (s) | 2.506 | 0.321 | [1.917, 3.123] |
| DAE | Bias | -0.001 | 0.045 | [-0.090, 0.086] |
|  | RMSE | 1.004 | 0.035 | [0.938, 1.076] |
|  | Time (s) | 0.312 | 0.038 | [0.283, 0.395] |
| Bayesian PCA | Bias | -0.000 | 0.043 | [-0.087, 0.084] |
|  | RMSE | 0.961 | 0.034 | [0.896, 1.033] |
|  | Time (s) | 0.180 | 0.023 | [0.150, 0.223] |
| Da Xu DL | Bias | -1.796 | 0.052 | [-1.893, -1.689] |
|  | RMSE | 2.189 | 0.054 | [2.082, 2.289] |
|  | Time (s) | 11.952 | 2.963 | [6.224, 17.006] |
| SoftImpute | Bias | -0.001 | 0.044 | [-0.087, 0.084] |
|  | RMSE | 0.994 | 0.039 | [0.921, 1.077] |
|  | Time (s) | 0.063 | 0.046 | [0.040, 0.207] |
| KNN | Bias | -0.005 | 0.057 | [-0.112, 0.108] |
|  | RMSE | 1.247 | 0.038 | [1.175, 1.320] |
|  | Time (s) | 0.104 | 0.022 | [0.095, 0.198] |

*Note:* Bias = mean difference between imputed and true values (closer to zero is better), RMSE = root mean squared error against known truth (lower is better), Time = execution time per iteration on complete cases (n=367). All values rounded to three decimal places. 95% CI calculated using percentile method.

**Figure S1.**
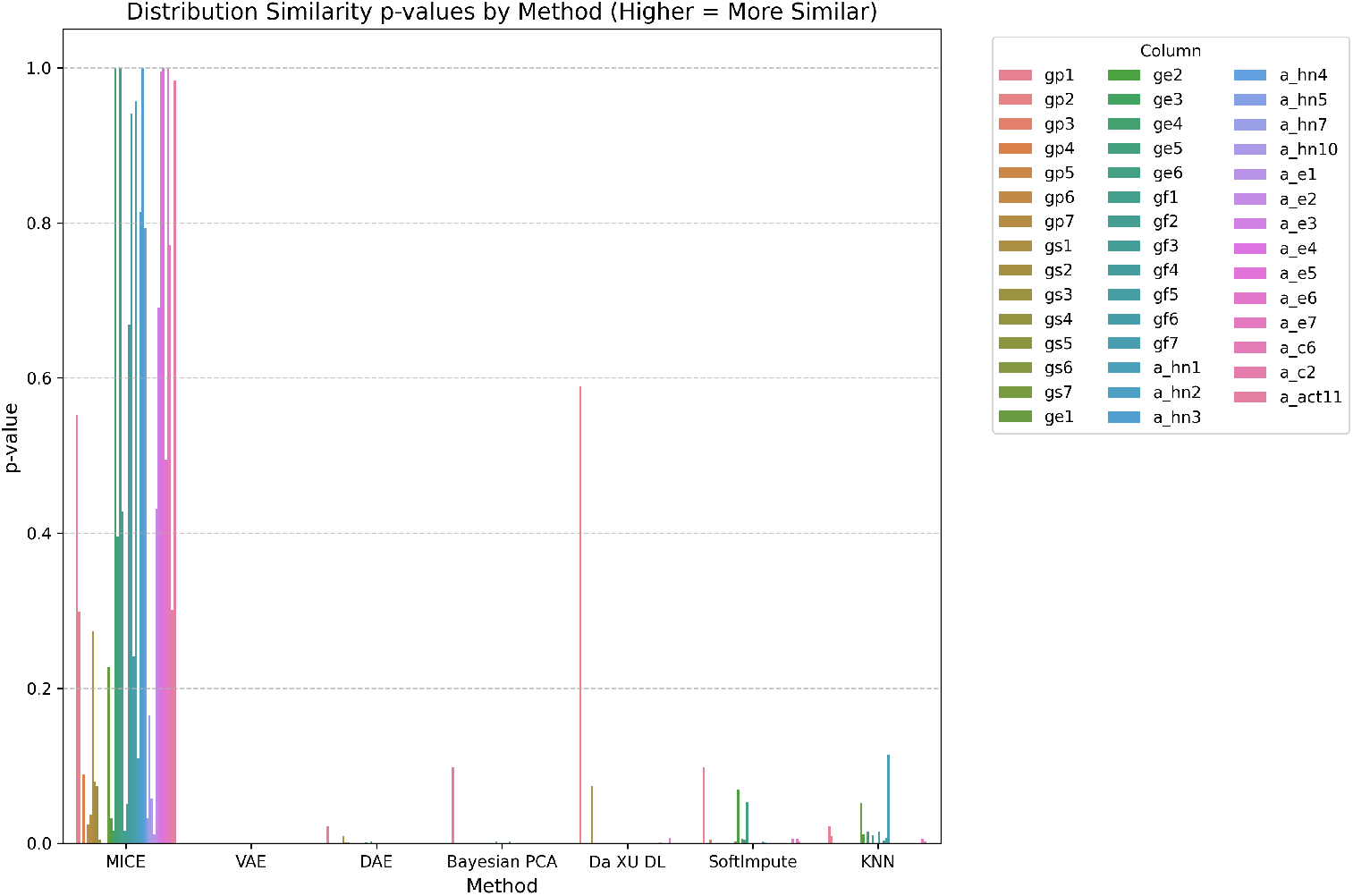
Distribution Similarity p-values by Imputation Method. Kolmogorov-Smirnov test p-values comparing observed versus imputed value distributions across 44 FACT-E variables (higher values indicate better distribution preservation). MICE demonstrated superior performance with many p-values above 0.05, indicating failure to reject the null hypothesis of identical distributions for multiple variables. Most other methods showed p-values near zero, suggesting significant distributional differences from observed data. Results complement the KS statistics in demonstrating MICE’s superior distribution preservation capabilities.

**Figure S2.**
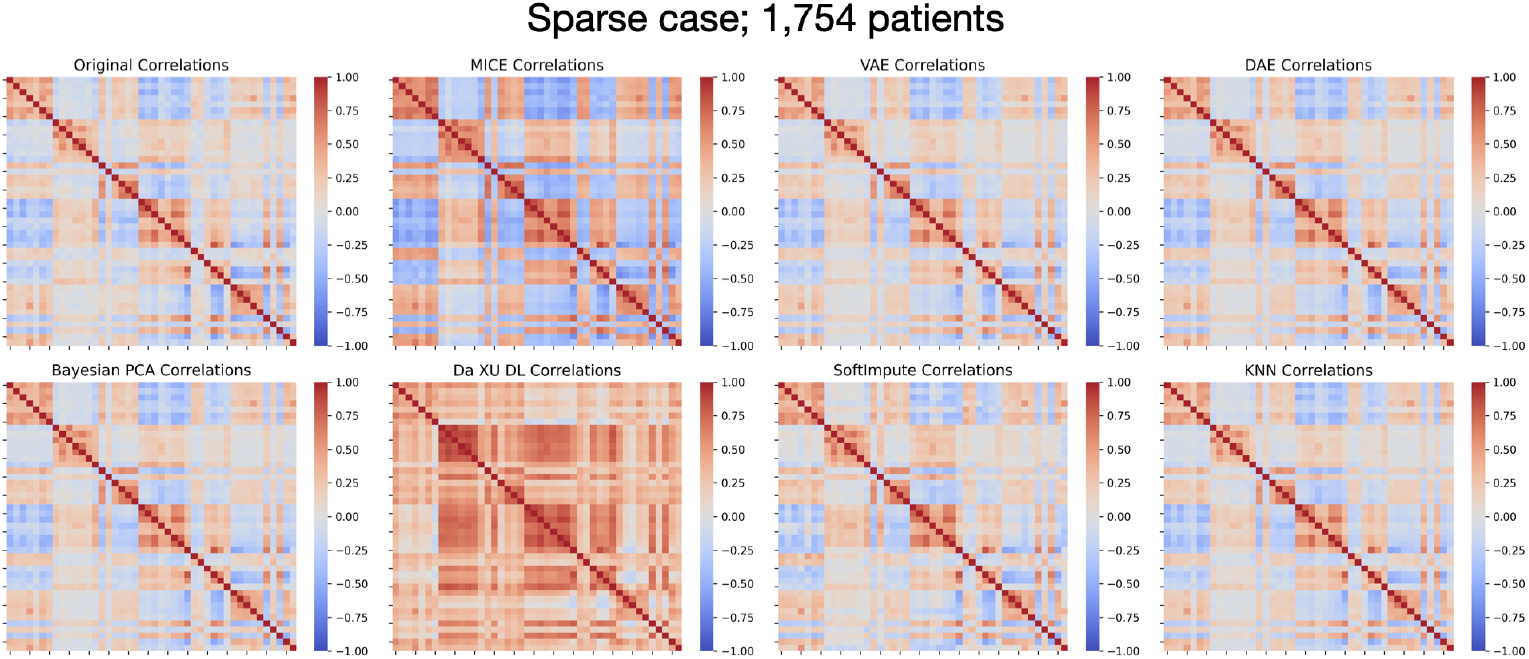
Correlation Matrix Across Imputed Results and Baseline Correlations in the More Sparse Data. Heatmaps comparing correlation structures between 44 FACT-E variables for original data (baseline correlations observed in complete cases) and each imputation method under increased missingness conditions (45.3-68.5%). Variable ordering is the same as Figure 5. Red indicates positive correlations, blue indicates negative correlations, with intensity representing correlation strength (-1 to +1). VAE, DAE, Bayesian PCA, and KNN maintained correlation patterns similar to the original structure, while SoftImpute showed moderate preservation. MICE exhibited notable deviations, and Da Xu et al. method^29;30^ introduced substantial artificial correlations (excessive red regions) not present in the original data, particularly affecting inter-domain relationships between FACT-E subscales.

**Figure S3.**
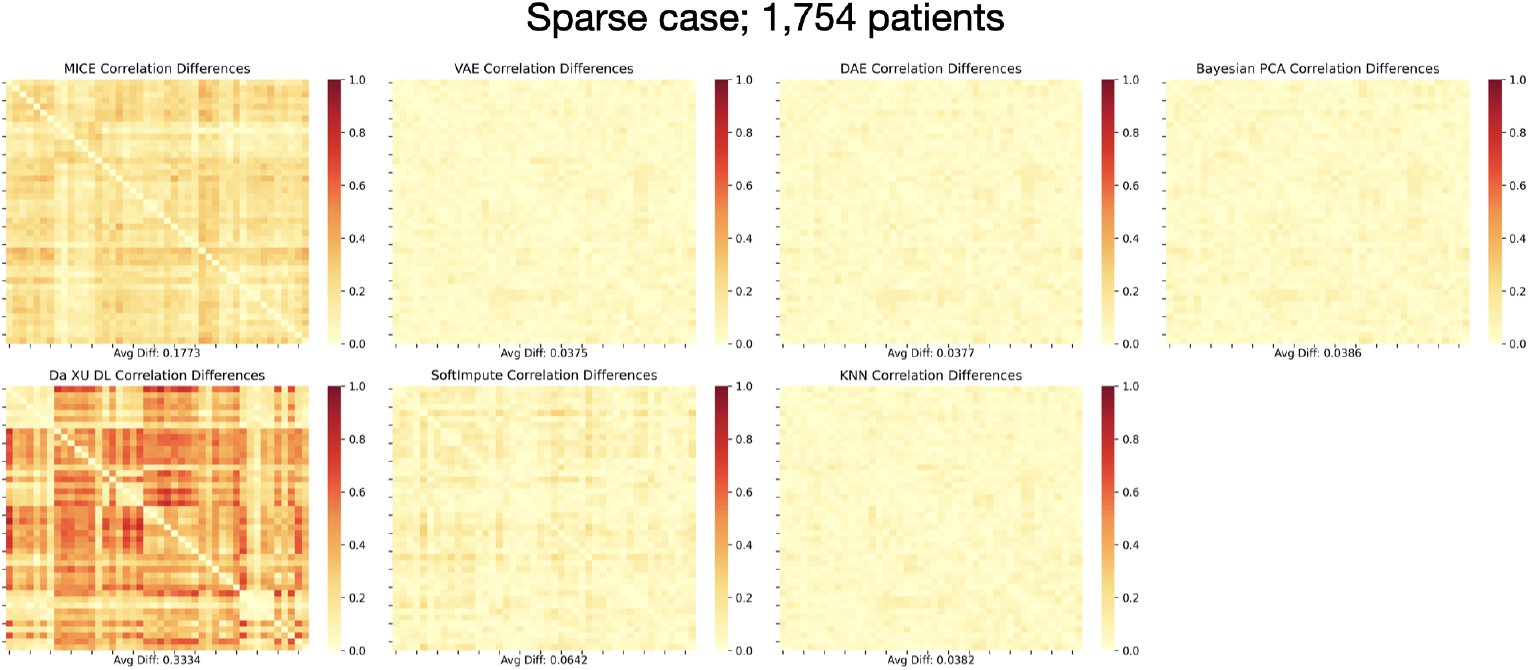
Absolute Correlation Differences Between More Sparse Data and Each Imputed Matrix. Absolute differences between original and imputed correlation matrices (lower values indicate better preservation) under challenging sparsity conditions. Variable ordering is the same as Figure 5. VAE (0.0375), DAE (0.0377), Bayesian PCA (0.0386), and KNN (0.0382) achieved exceptional preservation with minimal deviation from original patterns. SoftImpute (0.0642) showed moderate preservation, while MICE (0.1773) demonstrated degraded performance. Da Xu et al. method^29;30^ exhibited poor preservation (0.3334) with substantial correlation distortions across all variable pairs, indicating introduction of artificial relationships that compromise clinical interpretability of the quality-of-life structure.

## Footnotes

1 Variable labels follow the official FACT-E (Functional Assessment of Cancer Therapy-Esophageal) scoring guidelines. Complete variable definitions and scoring instructions are available in the FACT-E administration manual^25^ or at www.facit.org.

2 GS7 (“I am satisfied with my sex life”) exhibited the highest missingness rate, likely reflecting patient reluctance to respond to intimate personal questions in clinical settings.

## Notes

### Competing Interest Statement

The authors have declared no competing interest.

### Funding Statement

This research was funded with generous support from the Rossy Cancer Network - Cancer Quality and Innovation program.

### Author Declarations

Centre for Applied Ethics of McGill University Health Centre gave ethical approval for this work.

### Summary of Updates

The author list has been updated; The manuscript has undergone another round of review; The literature review and simulation study sections have been added, and the tables and figures have been updated accordingly.

